# Including non-client-facing provider time in calculating costs of health services

**DOI:** 10.1101/2025.06.04.25328992

**Authors:** Lori A Bollinger, Joseph Corlis

## Abstract

It is critical to calculate correctly the costs of implementing health interventions to generate reliable budgets, perform planning functions (including staff planning), and conduct economic evaluations. With the recent decrease in funding for international development assistance for health and the emphasis on country-led implementation of health programs in low- and middle-income countries, understanding the true cost of providing health services is even more important. As responsibility for funding and delivering health services shifts, inaccurate cost estimates can result in under-resourced disease response programs, ultimately lowering health outcomes. Costing studies, which aim to provide details on the financial and human resources necessary to deliver health services, rarely account for the time healthcare providers spend preparing for patient visits or following up with patients after a visit is complete. This deficiency in cost data is often acknowledged by researchers but seldom corrected, and consequently health policymakers do not know the true cost of staffing health facilities. In this paper, we present an updated methodology on how non-client-facing and non-clinical provider time should be incorporated into activity-based costing and management (ABC/M) applications. These methods are also applicable to traditional time-motion costing studies, including survey instrument changes and changes in calculating provider and operational costs. We present and discuss illustrative results which indicate that provider costs could increase by approximately 50% when non-client-facing provider time is included; operational costs could also increase, but likely by a smaller percentage. It is encouraging that we now have a simple, low-cost fix to the important issue of including provider costs correctly in both ABC/M and time-motion costing applications.

## Introduction

It is critical to calculate correctly the costs of implementing health interventions to generate reliable budgets, perform planning functions (including staff planning), and conduct economic evaluations. With the recent shifts in donor funding for health programs in low- and middle-income countries (LMICs) [1], emphasis on country-led implementation, financing, and sustainability planning [2] for health programming—including HIV; malaria; tuberculosis; reproductive, maternal, newborn, child, and adolescent health; and noncommunicable disease responses—has increased. As responsibility for providing these health services shifts to LMIC governments, understanding the true cost of providing health services is essential.

Health financing and economics research aims to provide this information for health policymakers and program planners. However, many studies that propose to demonstrate the costs of health services exclude a critical component: non-client-facing provider time, which is the time a provider spends at a health service site conducting activities that directly relates to providing health services while not directly interacting with a client [3 – 5]. Sometimes such non-client-facing provider time is critical to a single health care visit (e.g., charting, preparing for a procedure, preparing a prescription to be distributed, interpreting lab results) while other times provider time is spent on non-clinical work that cannot be attributed to a single health service type or visit (e.g., administration, participation in training).

Recently, the Activity-Based Costing and Management (ABC/M) methodology was endorsed and adopted by the United States President’s Emergency Plan for AIDS Relief (PEPFAR), the United States Agency for International Development (USAID), the United Nations Joint Programme on HIV/AIDS (UNAIDS), and other partners to estimate the costs of delivering HIV and other health services [6]. One aspect of this methodology, known as Time-Driven Activity-Based Costing (TDABC), relies on a data collector to shadow a client^1^ throughout a health care visit, recording the start and end times for each step in the service (e.g., registration, vitals taken, consultation). TDABC also requires that data collectors record other characteristics of the health care visit, including which staff members of the health service site interact with clients (e.g., receptionists, nurses, doctors, lab technicians) as well as any laboratory tests conducted, equipment used, medicines dispensed, and other supplies used or distributed in each step. One significant limitation of the current TDABC approach, which is widely acknowledged [7], is that data collectors are only able to record client-facing provider time (i.e., the time a provider spends with a client). Because data collectors shadow the client, they do not capture non-client-facing provider time or time spent on non-clinical care at the facility. This limitation results in significantly underestimating the time and resulting provider costs required to provide a health service, which in turn has important implications for planning and budgeting purposes. Note that this limitation also exists in costing studies that utilize a time-motion costing approach [8], which is a commonly used costing methodology that also tracks the time spent delivering care.

As part of the ABC/M methodology updating undertaken by the USAID-funded Analytics for Advancing the Financial Sustainability of the HIV/AIDS Response (AFS) project, we explored how non-client-facing and non-clinical provider time should be incorporated into the calculation of ABC/M costs. In the Materials and Methods section below, we present the underlying research and recommend a methodology that any costing study that uses time-motion calculations can apply. In the Results section, we present a practical application of the approach utilizing existing results in the published literature which illustrates the importance of including costs beyond client-facing provider time in calculations. Finally, the Discussion section describes the implications of applying the updated methodology, including planning and budgeting processes.

## Materials and Methods

### Materials

We searched both peer-reviewed and grey literature to identify approaches that have been used to account for non-client-facing and non-clinical provider time spent at a health facility or other service delivery site. While not exhaustive, our search identified three different approaches, each with respective pros and cons, which we drew from to develop our approach:

1. Brazil costing study [9]: In research undertaken in Brazil, a survey instrument was developed to measure the percentage of time spent by providers on various activities. The survey instrument was administered at the facility level (i.e., the questions pertain to the average values of the various cadres employed at each facility rather than to each individual provider). The survey instrument defined four main categories of provider time; note that these are oriented to the objective of the study, which is to examine non-clinical provider time in more detail:
  - time spent on providing clinical services
  - time spent on capacity building and mentoring
  - time spent on general administration and operations
  - time spent on monitoring and evaluation and health care management information systems.
2. American Medical Association (AMA) coding guidance [10]: Recently, in collaboration with the United States Centers for Medicare & Medicaid Services (CMS), the AMA revised its evaluation and management office visit documentation and coding requirements. The AMA provided guidance on these new rules, stating that physician or other qualified health professional time includes the following activities (when performed):
  - Preparing to see the patient (e.g., review of tests)
  - Obtaining and/or reviewing separately obtained history
  - Performing a medically necessary appropriate examination and/or evaluation
  - Counseling and educating the patient/family/caregiver
  - Ordering medications, tests, or procedures
  - Referring and communicating with other health care professionals (when not reported separately)
  - Documenting clinical information in the electronic or other health record
  - Independently interpreting results (not reported separately) and communicating results to the patient/family/caregiver
  - Care coordination (not reported separately) The AMA guidance states not to count time spent on the following:
  - The performance of other services that are reported separately
  - Travel
  - Teaching that is general and not limited to discussion that is required for the management of a specific patient
3. Published literature in family medicine [11, 12]: We identified two studies that measured family medicine physician time in the United States according to various categories. In the first study, physician time was disaggregated into four types of tasks:
  - face-to-face patient care
  - visit-specific work outside the exam room (e.g., updating medical records)
  - work related to patients not currently being seen (e.g., communicating about test results after the visit)
  - other tasks, including administrative tasks and mentoring.

The second study focused in a more disaggregated way on measuring non-client-facing time, listing multiple activities both related to clients and not related to clients.

Below, we apply aspects of the three approaches listed above to create the updated approach that we recommend for including non-client-facing and non-clinical provider time when calculating costs of delivering health services. First, we discuss how to categorize provider time by combining the approaches above, as well as the subsequent implications for the design of the survey instrument. Second, we discuss the implications of this updated approach in how costs are calculated, including how clinical provider costs obtained through the TDABC process are calculated and also how to incorporate the non-clinical provider costs into operational costs at the facility level.

We note that, in addition to incorporating methodological aspects from the third approach in the Methods section below, the Results section uses empirical results from the studies in calculating illustrative results.

### Methods

The first issue we considered when re-designing the survey instrument was how to categorize provider time. The approach used in the Brazil costing study, listed above, was designed to identify various aspects of non-clinical time, especially time spent on mentorship and research. While ABC/M applications do not require this level of detail in categorization, the approach of obtaining data at the facility level data was appealing. The AMA costing guidance provided some useful examples, particularly of specific clinical activities, but did not categorize the activities in a way that was helpful for ABC/M applications. In the approach drawn from family medicine studies, the first study categorized activities in a very useful way, and both studies in the third approach supplied good examples of activities contained within the categories. Thus, we utilize the categories from the first study of the third approach (see Table 1), and use examples from both family medicine studies as well as the AMA guidance approach to delineate which activities might be included in each category in the revised survey instrument:

**Table 1:** Revised categories for provider time.

| *Allocation of provider time spent on various activities in a typical day* |  |  |  |  |
| --- | --- | --- | --- | --- |
| % of time spent on client-facing activities | % of time spent on non-client-facing activities outside the exam room (current visit) | % of time spent on non-client-facing activities outside the exam room (post visit) | % of time spent on other activities (e.g., administrative, mentoring) | % time spent on lunches/breaks |

Sources: [9 – 12].

Our second issue in redesigning the time-motion data collection instrument was the level at which the instrument should be administered. While the family medicine approach, which included two studies detailing individual-level physician-reported time-motion information for providers, was appealing, the additional cost of interviewing each provider at each facility regarding their allocation of time would have been substantial, adding to an already-expensive costing exercise. We considered having the data collectors also observe a few providers from each cadre at each facility to assess their time-motion data in addition to observing the client, but we realized that even that extra cost would be prohibitive. Therefore, we developed our approach to administer a survey of key informants at the facility level that asks for each cadre’s time allocation for each facility, but using the new categories as shown in Table 1.

Utilizing the data obtained by applying this updated approach has significant implications for how provider and operational ABC/M costs need to be calculated in several ways.^2^ In prior applications, the capacity cost rate (CCR) for providers included client-facing clinical time only (i.e., it excluded consideration of both non-client-facing clinical time and non-clinical time at the facility) [13, 14].

Thus, the first change in our updated methodology is to consider both non-client-facing clinical time and non-clinical time at the facility when calculating the CCR for providers. The CCR is calculated based on the total number of minutes a provider has available during the year (see Table 2 for an illustrative example). The calculation of the number of working days per year remains the same as before, where weekends, paid time off (including vacation, sick, and holidays), and other professional activities not facility-related (e.g., conferences) are excluded when calculating available days. In the illustrative example below, the net result of subtracting these days is a total of 208 working days per year. The calculation of available clinic hours, however, is slightly different than previous CCR calculations. Here, other (non-clinical) time is included in the number of hours available to a provider in a day, instead of just clinical time; previously ignored, the costs of this non-clinical time are included here in operational costs at the clinic level. We note that including non-clinical time in the available clinic hours per day will lower the CCR slightly relative to previous results.

**Table 2:** Illustrative example of a capacity cost rate calculation for provider time.

| **Available Days** |  |
| --- | --- |
| Available days per year | 365 |
| Less Weekends (days) | 104 |
| Less vacation, holidays, sick (days) | 28 |
| Less training, conferences, research, continuing education (days) | 25 |
| *Total working days per year* | *208* |
| **Available Facility Hours per day** |  |
| Time spent at facility (hrs./day) | 9.0 |
| Less time for breaks, lunch, etc. (hrs./day) | 1.0 |
| *Total available clinical time (client-facing time, non-client-facing time during current visit, post-visit non-client-facing time) and other time (e.g., administrative, mentoring) (hrs./day)* | *8.0* |
| **Capacity Cost Rate** |  |
| Capacity per year (208 days * 8.0 hours/day = 1,664 hours) | 99,840 minutes |
| Total annual compensation (salary + benefits) | USD \$5,763 |
| *Capacity Cost Rate (USD 5,763/99,840 minutes)* | *USD \$0.06/minute* |

Source: [7].

After the above adjustment is made in calculating the CCR, the newly revised CCR must then be applied correctly. Because data collection during TDABC centers on following the client while health services are being provided, only recording the time a client spends with a provider, (i.e., the client-facing provider time), the direct service delivery portion of the cost (i.e., excluding operational costs) must be adjusted to include the non-client-facing provider time (i.e., both the non-client-facing time expended on the day of the current visit and the non-client-facing time that occurs on days after the visit). In addition, the non-clinical provider time must be included in operational costs at the facility level utilizing the recorded client-facing provider time. The equation below represents available clinic hours per day from Table 2; note that only observations on client-facing time (**bolded**) are available via TDABC:

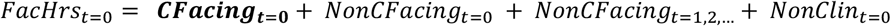

where:

- *FacHrs*_*t*=0_ is the total available provider time per day at the facility on the day of the visit (t=0);
- ***CFacting***_***t***=**0**_ is the client-facing time spent by the provider on the day of the visit (t=0) as observed via TDABC;
- *NonCFacing*_*t*=0_ is the non-client-facing time spent by the provider on the day of the visit (t=0), e.g., medical record keeping, ordering lab tests;
- *NonCFacing*_*t*=1,2,…_ is the non-client-facing time spent by the provider after the day of the visit (t=1, 2, …), e.g., reporting back lab tests results; and
- *NonClin*_*t*=0_ is the non-clinic provider time on the day of the visit (t=0), e.g., administration.

While we only know the number of minutes spent on client-facing time from TDABC results, we do know the percentage of time spent on the four components of the right-hand side of the equation above based on facility-level data captured by the survey instrument. Thus, we can calculate a scaling factor to adjust the client-facing minutes by the appropriate percentage(s) to calculate total clinical provider time:

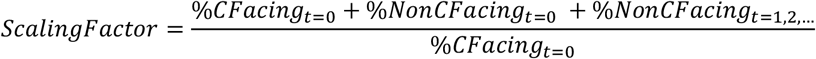

where:

- *ScalingFactor* is the (time-invariant) scaling factor to be applied;
- %*CFacting*_*t*=0_ is the percentage of client-facing time spent by the provider on the day of the visit (t=0) as observed via TDABC;
- %*NonCFacing*_*t*=0_ is the percentage of non-client-facing time spent by the provider on the day of the visit (t=0); and
- %*NonCFacing*_*t*=1,2,…_ is the percentage of non-client-facing time spent by the provider after the day of the visit (t=1, 2, …).

This scaling factor is then applied to the number of minutes derived from TDABC to calculate the total amount of time a provider spends on clinical activities. This number is then multiplied by the (correctly updated) CCR to calculate the total cost of the clinical time spent by providers.

The costs of non-clinical time (e.g., administrative tasks) are included in the operational costs at the facility level. They can be calculated directly by multiplying the percentage of time spent on non-clinical activities, per Table 2, by the cadre’s salary for each cadre at the facility; the costs are then added to the facility-level operational costs (see Results section below for an illustration of these calculations).

## Results

In this section, to provide illustrative results, we apply the time allocation for provider tasks described above [11] to recent ABC/M costing results from Tanzania [13, 15] in order to calculate illustrative updated provider and operational costs.^3^ Table 3 displays the hours associated with the revised categories for provider time as well as the relevant percentages. Note that, because the time spent on lunches and breaks was not recorded in the study, and the assumption is required to be able to calculate operational costs correctly, we assume lunches and breaks take up one hour per day:

**Table 3:** A recalculation of the number of hours (% of total) spent by a provider on facility activities.

| Time spent on client-facing activities in hours (% of total) | Time spent on non-client-facing activities outside the exam room (current visit) in hours (% of total) | Time spent on non-client-facing activities outside the exam room (post visit) in hours (% of total) | Time spent on other activities (e.g., administrative, mentoring) in hours (% of total) | Time spent on lunches/breaks in hours (% of total) |
| --- | --- | --- | --- | --- |
| 5.9 (56%) | 1.25 (12%) | 1.97 (19%) | 0.33 (3%) | 1.0 (10%) |

Source: [11].

Based on the percentages calculated in Table 3, the scaling factor for clinical activities for a provider equals 154.6%.^4^ We apply this to the original Tanzania ABC/M costing results, displayed in column (B) of Table 4, to calculate updated provider costs, displayed in column (D).

**Table 4:** Service line costs per visit and variation across facilities in Tanzania (USD).

| (A)<br>Service line | Original |  | Updated |  | (F)<br>% change in<br>mean cost<br>based on<br>recalculation |
| --- | --- | --- | --- | --- | --- |
|  | (B)<br>Mean cost | (C)<br>% of total | (D)<br>Mean cost | (E)<br>% of total |  |
| *ART—Stable\** | *\$90.88* | *100.0%* | *\$92.72* | *100.0%* | *2.0%* |
| Providers | \$3.00 | 3.3% | \$4.64 | 5.0% | 54.6% |
| Space/equipment | \$0.40 | 0.4% | \$0.40 | 0.4% | |
| Operational | \$1.56 | 1.7% | \$1.72 | 1.9% | 10.3% |
| Consumables | \$85.96 | 94.6% | \$85.96 | 92.7% | |
| *ART—Unstable\** | *\$178.32* | *100.0%* | *\$184.02* | *100.0%* | *3.2%* |
| Providers | \$9.72 | 5.5% | \$15.02 | 8.2% | 54.6% |
| Space/equipment | \$1.44 | 0.8% | \$1.44 | 0.8% | |
| Operational | \$4.32 | 2.4% | \$4.84 | 2.6% | 12.0% |
| Consumables | \$162.72 | 91.3% | \$162.72 | 88.4% | |
| *HTC* | *\$3.67* | *100.0%* | *\$4.16* | *100.0%* | *13.4%* |
| Providers | \$0.82 | 22.3% | \$1.27 | 30.5% | 54.6% |
| Space/equipment | \$0.11 | 3.0% | \$0.11 | 2.6% | |
| Operational | \$0.50 | 13.6% | \$0.54 | 13.1% | 8.7% |
| Consumables | \$2.24 | 61.0% | \$2.24 | 53.8% | |

<sup>4</sup> The calculation is: $(56\% + 12\% + 19\%) / 56\% = 154.6\%$ .
<sup>5</sup> As an example, the updated provider cost (column D of Table 4) for a stable ART client is \$4.64, accounting for 87% of the full cost of a provider. We multiply by $100\% / 87\%$ to calculate the full cost of \$5.33. We then multiply this by 3% (\$0.16), which is then added to the original cost (\$1.56 in column B) for “ART-Stable” to calculate the updated operational costs (\$1.72 in column D).

|  |  |  |  |  |  |
| --- | --- | --- | --- | --- | --- |
| *PMTCT* | \$22.12 | 100.0% | \$22.82 | 100.0% | 3.2% |
| Providers | \$1.16 | 5.2% | \$1.79 | 7.9% | 54.6% |
| Space/equipment | \$0.13 | 0.6% | \$0.13 | 0.6% | |
| Operational | \$0.44 | 2.0% | \$0.50 | 2.2% | 14.1% |
| Consumables | \$20.40 | 92.2% | \$20.40 | 89.4% | |
| *PrEP* | \$6.77 | 100.0% | \$7.12 | 100.0% | 5.2% |
| Providers | \$0.59 | 8.7% | \$0.91 | 12.8% | 54.6% |
| Space/equipment | \$0.04 | 0.6% | \$0.04 | 0.6% | |
| Operational | \$0.15 | 2.2% | \$0.18 | 2.5% | 21.0% |
| Consumables | \$5.99 | 88.5% | \$5.99 | 84.1% | |
| *VMMC* | \$28.00 | 100.0% | \$30.38 | 100.0% | 8.5% |
| Providers | \$3.97 | 14.2% | \$6.14 | 20.2% | 54.6% |
| Space/equipment | \$0.43 | 1.5% | \$0.43 | 1.4% | |
| Operational | \$0.91 | 3.3% | \$1.12 | 3.7% | 23.3% |
| Consumables | \$22.69 | 81.0% | \$22.69 | 74.7% | |

ART=antiretroviral therapy, HTC=HIV testing and counseling, PMTCT=prevention of mother-to-child transmission, PrEP=pre-exposure prophylaxis, VMMC=voluntary medical male circumcision
\*Annualized cost presented for ART-Stable and ART-Unstable. All other conditions presented as cost per encounter
Sources: Columns A-C, [13, 15]; columns D-F, authors' calculations.

Further, because we do not have the underlying study data to calculate revised operational costs, for our illustrative example here we use the results from above to calculate the impact of including non-clinical provider time in operational costs. Again, this won’t be necessary when implementing the updated methodology in future ABC/M applications, as the percentage of non-clinical time for each cadre will be obtained in the updated facility survey. Here, we use the results above to calculate updated operational costs. We know the updated cost of clinical activities for each intervention, and from Table 3 we know that it is 87% of the total provider time. We also know from Table 3 that non-clinical time is 3% of the total provider time. Using this relationship, we interpolate and add the resulting costs to the operational costs for each intervention.^5^

The overall impact of updating both provider costs and operational costs ranges from a low of 2.0% for stable antiretroviral therapy clients (“ART – Stable”), which increases from $90.99 to $92.72 per client per year, to a high of 13.4% for HIV testing and counseling (HTC) clients, which increases from $3.67 to $4.16 per client per facility visit (see column F in Table 4). The results indicate that the impact is relatively lower, ranging between 2.0% and 5.2%, when antiretrovirals (ARVs) are being provided; for example, for “ART – Stable” clients (2.0%), for “ART – Unstable” clients (3.2%), for clients of prevention of mother-to-child transmission (PMTCT) services (3.2%), and for pre-exposure prophylaxis (PrEP) clients (5.2%). The other two interventions exhibit higher relative increases, showing an 8.5% increase for voluntary medical male circumcision (VMMC) clients ($28.00 to $30.38) and a 13.4% increase for HTC clients ($3.67 to $4.16). We note that the procedure kits used in VMMC services, while not as expensive as ARVs, account for a large overall percentage of VMMC costs.

Delving into the details, the results indicate that the disaggregated provider costs all increase by the same relative amount: 54.6%. This is because the scaling factor is a relative (percentage) factor and is applied directly to the original provider costs. However, the extent to which the absolute amount changes, which is illustrated by the share of providers’ cost in the total cost of the intervention, varies by the relative importance of provider costs in the total cost. For example, for “ART – Stable” clients, which has the lowest provider cost share in the original costs (3.3%) and exhibits the lowest increase overall, the share accounted for by providers only increases by 1.7 percentage points (3.3% to 5.0%) in the overall updated cost. For HTC clients, which has the highest initial provider cost share (22.3%) and highest increase overall, the share accounted for by providers increases by 7.2 percentage points (22.3% to 30.5%) in the overall updated cost.

The impact of the updated methodology on operational costs exhibits a different pattern, as the updated costs are based on absolute changes rather than the relative changes used to update the provider costs. The absolute levels of all operational costs increase, ranging from a low of $0.03 for updated PrEP costs per client to a high of $0.52 for updated “ART – Unstable” costs per client. However, the pattern of changes in the share of operational costs is different. All interventions delivering ARVs in some form— “ART – Stable,” “ART – Unstable,” “PMTCT,” and “PrEP”—exhibit a change of 0.2 percentage points in the share of operational costs in overall costs, even when the changes in the absolute amounts differ. The other two interventions actually decrease in the share of operational costs in overall mean costs, with updated operational costs contributing 0.1 percentage points less to total updated VMMC costs per client and contributing 0.5 percentage points less to total updated HTC cost per client.

## Discussion

We combined various aspects of the existing costing literature to update the ABC/M costing methodology to include consideration of non-client-facing provider time in both provider and operational costs. There are three key findings from this analysis. First, illustrative results indicate that provider costs increase by approximately 50% when non-client-facing provider time is included in HIV services provided in Tanzania. This has significant implications for budgeting and planning; increasing personnel costs by 50% is critical to include for annual operational budgeting as well as longer-term planning, especially as countries assume greater responsibility for health programming and financing. Perhaps more importantly, it is also critical to include consideration of the extra time required by providers in care provision when constructing staffing plans. Based on our updated calculations for Tanzania, we would expect that, if the original ABC/M results were used in staffing plans, the services included in the study faced an underestimation of two-thirds the required staff to conduct all client visits. This could contribute to the long wait times for health services that Tanzania’s ABC/M study—and many other studies—have identified [16 – 19].

Second, operational costs also increase in our illustrative example when non-clinical time at the facility is included. The impact here is not as significant as the increases in provider costs, but the additional costs are no less critical for policymakers and health program managers to have for health service plans and evaluations. Further, as discussed below in limitations, the illustrative data used here—based on family medicine provider allocation in the United States, most of whom have medical assistants to perform administrative tasks—may bias downwards the additional amount contributed to operational costs using the updated methodology in LMICs where different providers must perform those tasks.

Third, we should note that, while the impact of the updated methodology is certainly important here in our illustrative example using the delivery of HIV services, it will likely be even more important for other health services for which the commodities’ share of the total mean cost is lower (e.g., antenatal care).

There are some limitations to this analysis. One limitation is that we are not using data we collected to demonstrate the impact of the updated methodology on costing study calculations. Instead, we have combined results that use the original methodology with various assumptions to calculate updated results. We believe the results are reliable enough for the sake of argument (e.g., a 50% increase in provider time due to time spent on non-client-facing service provision is reasonable) but the actual impact of our updated methodology will depend on the empirical results obtained when it is implemented in the field.

A second limitation is that, again for illustrative purposes, we have used time allocation figures based on family medicine provider-reported time in the United States. As noted above, these providers likely have medical assistants who assist in the various tasks, particularly administrative tasks, so updated costs based on these time allocation figures may be understated, particularly the updated operational costs. By applying the survey instrument to capture locally relevant time allocation percentages, we expect the scaling factor to be accurate.

Finally, as noted above as well, the cost components of HIV service provision may not be representative of cost components of other health services; in particular, the commodities cost for interventions supplying ARVs accounts for a large percentage of total intervention costs. When commodities represent a lower portion of total intervention costs, resulting in a relatively higher portion of provider costs in total intervention costs, using this updated methodology becomes even more important.

These limitations are, however, specific to this illustrative analysis, and disappear once the updated methodology is implemented in new ABC/M applications. It is encouraging that we now have a simple, low-cost fix to the important issue of including provider costs correctly in ABC/M and other time-motion costing applications; researchers can address this longstanding gap in costing information by adding a few fields to the facility-level interview questionnaire and adjusting the formulae used in calculations of provider and operational costs.

## Data Availability

All data produced in the present study are available upon reasonable request to the authors.

## Acknowledgements

Some of the research underlying this paper was supported by the United States Agency for International Development. The authors gratefully acknowledge comments received from Elan Reuben, Rebecca Ross, and the Global Technical Committee on ABC/M.

## Footnotes

1 We use the term *client* to indicate anyone who receives a health service or product; in many contexts, such individuals may also be referred to as a *patient, beneficiary, service user, end user*, or other similar term.

2 For resources that are used more than once, ABC/M requires multiplying a capacity cost rate (CCR), which is a cost per minute to use a resource, by the number of minutes that resource is used (e.g., provider time, laboratory equipment, furniture) to calculate costs [7].

3 Future ABC/M applications will, of course, record the time allocation for provider tasks for each cadre for each facility as part of their data-gathering efforts, so operational costs will be calculated using these obtained data.

4 The calculation is: (56% + 12% + 19%) / 56% = 154.6%.

5 As an example, the updated provider cost (column D of Table 4) for a stable ART client is $4.64, accounting for 87% of the full cost of a provider. We multiply by 100% / 87% to calculate the full cost of $5.33. We then multiply this by 3% ($0.16), which is then added to the original cost ($1.56 in column B) for “ART-Stable” to calculate the updated operational costs ($1.72 in column D).

